# Digital Health Tools Applications in Frail Older Adults - A Review Article

**DOI:** 10.1101/2024.02.01.24302134

**Authors:** Natthanaphop Isaradech, Wachiranun Sirikul

## Abstract

**Background:** Frailty is a common degenerative condition highly prevalent in adults over 60 years old. A frail person has a higher risk of morbidities and mortality when exposed to health-related stressors. However, frailty is a reversible state when it is early diagnosed. Studies have shown that frail people who participated in an exercise prescription have a greater chance to transition from frail to fit. Additionally, with a rapid advancement of technology, a vast majority of studies are supporting evidence regarding the digital health tools application on frail population in recent years.

**Objectives:** This review comprehensively summarizes and discusses about technology application in frail persons to capture the current knowledge gaps and propose future research directions to support additional research in this field.

**Methods:** We used PubMed to search literature (2012-2023) with pre-specified terms. Studies required older adults (≥40 years) using digital tools for frailty comparison, association, or prediction and we excluded non-English studies and those lacking frailty comparison or digital tool use.

**Findings:** Our review found potential etiognostic factors in trunk, gait, upper-extremity, and physical activity parameters for diagnosing frailty using digital tools in older adults.

**Conclusion:** Studies suggest exercise improves frailty status, emphasizing the need for integrated therapeutic platforms and personalized prevention recommendations.

## Introduction

Aging society is an inevitable ongoing trend in in the world. World Health Organization reported that within 2030 1 in 6 people in the world will be aged over 60 years and by 2050 number of persons aged over 60 years old is expected to reach 426 million (1,2). This trend is common in many countries and is attributed to a combination of factors such as improved healthcare and advancements in medical technology, which have allowed people to live longer (3). The world public health is now facing the challenges and opportunities that come with an aging society since older adults are linked to increased multiple chronic diseases, comorbidities and mortality (4–6).

Frailty is a clinical condition highly prevalent in the aged population in which a frail individual is more vulnerable to health-related risk exposure (7,8). Studies showed that this condition has been linked to increased hospitalizations, Emergency Department (ED) visits (9–12), poorer quality of life (13), impaired cognitive function (14), increased morbidity and mortality (15). Frailty is commonly defined by Fried et. al. 2001 using unintentional weight loss, gait speed, exhaustion, grip strength and physical activity as a clinical diagnostic criteria (16).

There has been an increase in studies on frailty in recent years since frailty could be decreased or reversed with a long-term-based exercise intervention (17,18). Fairhall et. al. 2014 conducted an randomized controlled trial of 241 community-dwelling older adults in Australia where the findings showed that exercise and nutrition intervention could significantly improve frailty status in the treatment group (19). The result agreed with Nakamura et. al. 2022 where 111 community-dwelling older people in Japan were randomly assigned to perform a home-based training during Covid-19 pandemic (20).

Several studies have employed digital tools to help diagnose and treat frailty as technology has improved and become more accessible (21,22). We believe that information technology can help us recognize frailty earlier, and that the earlier we identify this condition, the better healthcare providers can treat the patient with a better prognosis and health outcomes. This review aims to identify and summarize prospective characteristics, diagnostic models, and therapeutic studies in utilizing digital health technologies in community-dwelling frail older persons.

## Search Strategy

We used PubMed as our main source of published literature for our search strategy. The combinations of search terms were (“frailty*” OR “frail” OR “frail elderly”) AND (“digital” OR “machine learning” OR “smartphone*” OR “AI” OR “artificial intelligence” OR “deep learning” OR “device*”) AND (“older adults” OR “elderly” OR “elder” OR “old”). The selected publications in our review were limited to English publications and publications within 2012-2023. Additional literature found in systemic reviews and meta-analysis were manually selected to include in this review. Inclusion Criteria: 1.) The study recruited older adults aged at least over 40 years old; 2.) The study applied digital health tools to find association, causal relationship or make prediction between frail and non-frail population. Exclusion Criteria: 1.) The study was not written in English; The study did not demonstrate a comparison of result between frail and non-frail population; The study did not utilize digital tools.

## Characteristics studies

We found 9 relevant frailty characteristics studies and summarized them into Table 1. Most studies used various types of digital sensors to measure surrogate outcome of frailty and are categorized into 1.) Trunk parameter 2.) Gait parameter and 3.) Non-gait parameters

**Table 1.**
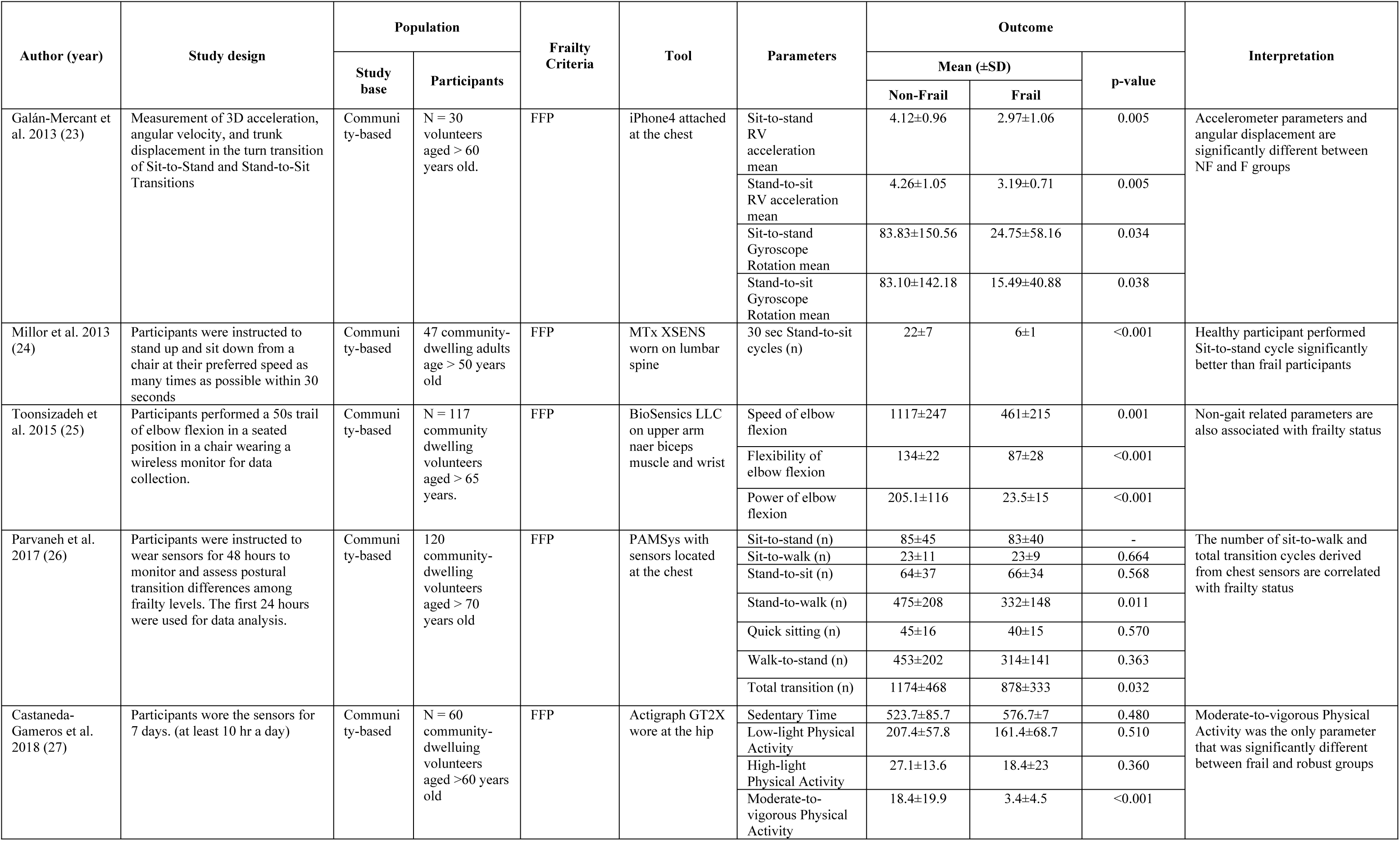
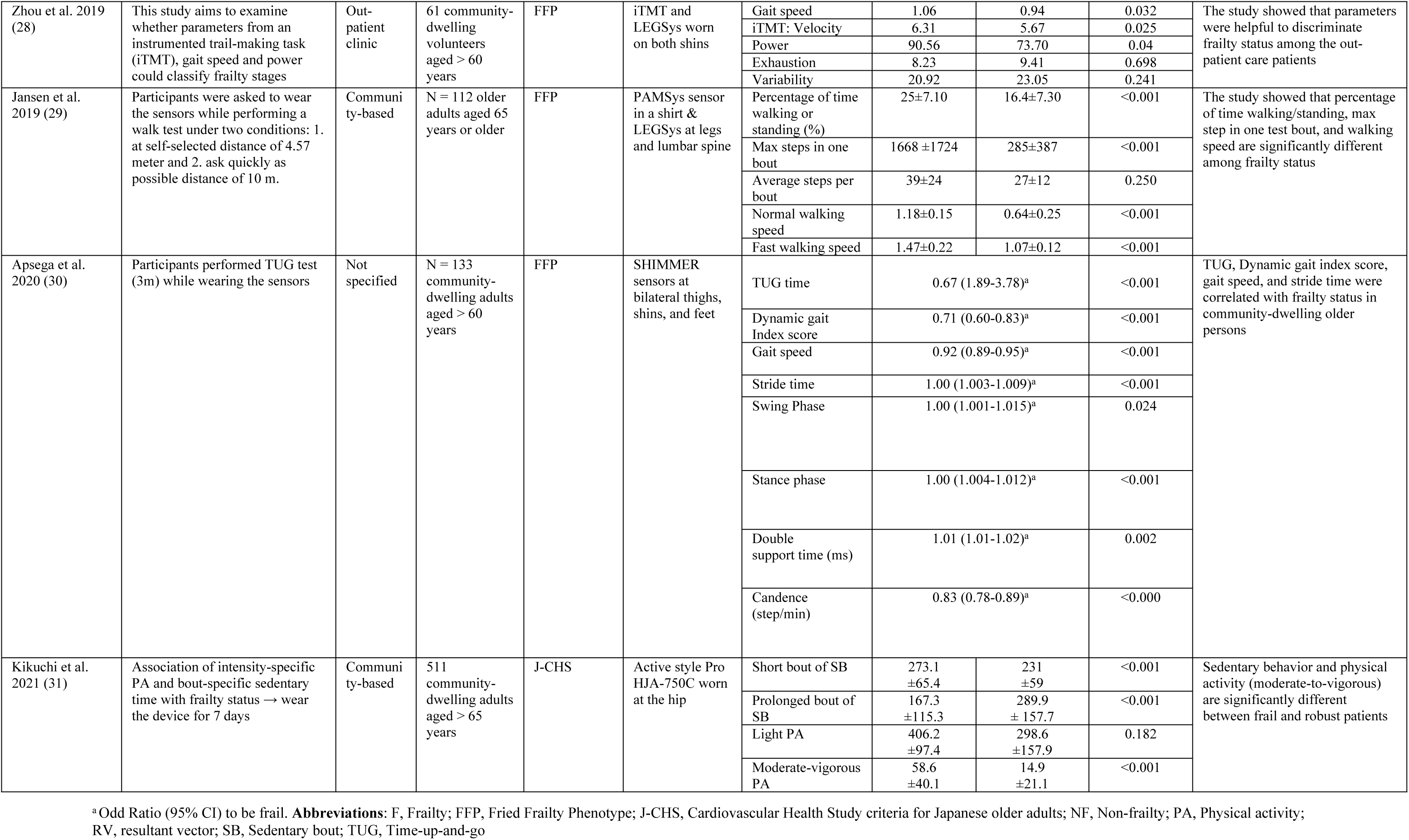
Characteristics Studies.

## Trunk parameter

Most studies’ methods involved researchers instructing volunteers to perform physical function tests while wearing a digital sensor that measures characteristics that likely represents frailty. Galan-Mercant et. al. (2013) studied 30 community-dwelling volunteers over the age of 60 who performed a sit-to-stand, stand-to-sit test while wearing an iPhone4 attached to the chest to assess 3D acceleration, angular velocity, and trunk displacement during the turn transition (23). The findings revealed all factors differed significantly between frail and non-frail subjects.

Parvaneh et. al. 2017 conducted a study using a wearable necklace-like sensors located at the chest of 120 community-dwelling participants aged over 70 years old to monitor and assess postural transition differences among frailty levels for 24 hours, and the results showed that the number of Stand-to-walk and total postural transitions were significantly different between groups (26). Millor et. al. (2013) asked 47 community-dwelling volunteers over the age of 50 to perform stand-up and sit-down from a chair as many times as they could in 30 seconds while wearing an inertial orientation tracking sensors on their lumbar spine (24). The study showed that healthy participants outperformed frail people with less sway on the sit-to-stand cycle.

Therefore, the parameters derived from sensors attached to the trunk such as 3D acceleration, velocity and postural sway while doing physical function tests could discriminate frail and robust in the community-dwelling older adults.

## Gait Parameter

Gait assessment was another method used by researchers to analyze diagnostic variables in frail older adults. Zhou et. al. 2019 investigated whether parameters from an instrumented trail making task (iTMT) and gait sensors worn on both shins to measure gait speed and iTMT derived parameters could distinguish between frail and robust participants (28). The findings revealed that gait speed and iTMT velocity were significant parameters that could help classify frailty status among the outpatient care population. Moreover, Jasen et. al. 2019 carried out an intervention research which 112 community-dwelling older persons were requested to wear a wearable sensor in a shirt while undertaking a walking test under two conditions: 1.) Walk a distance of 4.57 meters at your own speed; and 2.) Walking a 10-meter distance as rapidly as possible (29). The findings correlated with the previous studies, which suggested that the proportion of time spent walking and standing, the maximum steps in one test bout, and walking speed might all be potential predictors of frailty classification (30).

## Non-gait Parameters

To determine frailty status, other variables could be used in addition to those mentioned above. Toosizadeh et. al. (2015) studied the association between frailty status and non-gait parameters using a wearable gyroscope sensor attached to the upper arm and wrist of 117 community-dwelling adults over 65 years old to measure elbow function while performing a 50-second trail of elbow flexion in a seated position (25). The results revealed that the speed of elbow flexion, flexibility, and power of elbow flexion differed significantly between robust and frail participants. In the study by Castaneda-Gameros et. al. (2018), moderate-to-vigorous physical activity measured by a sensor that records acceleration and gyroscopic data worn on the hip for 7 days was associated with frailty status in community-dwelling old adults (27). Additionally, Kikuchi et. al. 2020 found the association of intensity-specific physical activity. The results showed that sedentary behavior and physical activity (moderate-to-vigorous) were significantly different between frail and robust in 511 Japanese community-dwelling participants aged over 65 (31).

As a result of the mentioned studies, there are multiple potential variables that could represent characteristics of frailty. Non-gait parameters appeared to have the highest clinical feasibility if researchers could integrate a model into a smartwatch since a wrist-worn device is simple to use and most older adults are already accustomed to wearing a smartwatch.

## Diagnostic Studies

Frailty identification is a clinically relevant topic since it is a condition that may be reversed from frail to robust. Several studies are being conducted to develop tools and diagnostic models for classifying frail and non-frail older adults. According to the authors’ evaluation of the published evidence in this field, there are two types of frailty diagnostic tools that use technology: 1.) Clinical Data; and 2.) Data derived from wearable devices and biological sensors which are summarized in Table 2.

**Table 1.** Diagnostic studies.

| Author (year) | Study design | Population |  | Predictors | Frailty-criteria | Tool | Model | Diagnosis performances | Interpretation |
| --- | --- | --- | --- | --- | --- | --- | --- | --- | --- |
|  |  | Study base | Participants |  |  |  |  |  |  |
| Prediction Model |  |  |  |  |  |  |  |  |  |
| Aznar-Tortonda et al. 2020 (32) | Cross-sectional observational study using an application for Android for data collection | Community-based | 621 older adults | Sex, age, polypharmacy, hospital admission in the last year, and diabetes. | FFP | Android Application | LGR | AUC 0.78 | Simple clinical history could be used for frailty classification in older adults |
| Sajeev et al. 2022 (33) | Cross-sectional Observational study (Development and internal validation with test sample) | Community-based | 656 independent community-dwelling adults aged 40-75 years old | 63 anthropometric, environmental, social, lifestyle and physiologic variables | CFS and FFP | In-person Health Assessment | LGR | AUC 0.69 | Machine learning methods are useful for frailty diagnosis however some variables might be hard to implement in clinical practice |
|  |  |  |  |  |  |  | LDA | AUC 0.69 |  |
|  |  |  |  |  |  |  | SVM | AUC 0.69 |  |
|  |  |  |  |  |  |  | RF | AUC 0.71 |  |
| Data derived from wearable devices and biological sensors. |  |  |  |  |  |  |  |  |  |
| Greene et al. 2014 (34) | Development of classifier models to assess frailty status using sensor-derived features of TUG, Five Time Sit to Stand and Balance tests | Community-based | 124 community dwelling older adults (mean age 75.9 ± 6.6 years, 91 female). 66 F, 58 NF | Time up and go test, Balance test, Five Time Sit to Stand | FFP | SHIMMER sensor worn on each shin, right thigh, L5 spine and sternum.<br>A pressure sensor for balance data. | SVM | Sensitivity 88.63%<br>Specificity 85.06% | TUG, FTSS and Balance test are good predictors for frailty classification using an SVM model |
| Schwenk et al. 2015 [35] | Participant walked 4.57 m in their home at self-selected speed. Balance was assessed during 15s quiet standing with feet together, eyes closed. PA | Community-based | N = 125 community-dwelling volunteers aged > 65 years old | Stride length, Double support, Balance parameters | FFP | LEGSSys, BalanSens, PAMSys with sensors located at shanks, thighs, and lumbar spine | LGR: NF and PF classification | AUC 0.86 | Gait parameters had the best performance to separate NF from PF and PF from F in aged-adjusted model |
|  |  |  |  |  |  |  | LGR: PF and F classification | AUC 0.84 |  |
| Tooiszadeh et al. 2015 (35) | Participants performed two 15s balance mechanisms between NF, PF and F individuals | Community-based | 122 older adults aged > 65 years old | Postural sway, age, BMI, OLCL parameters | FFP | BalanSens at lumbar spine and shin | LGR: Postural sway, age, and BMI | EO: Sensitivity 74%, Specificity 93%<br>EC: Sensitivity 74%, Specificity 83% | Body sway (and age/BMI), OLCL (and age/BMI) can be used for frailty screening tool (high sensitivity) |
|  |  |  |  |  |  |  | LGR: OLCL, age and BMI | EO: Sensitivity 94%, Specificity 98% |  |
|  |  |  |  |  |  |  |  | EC: Sensitivity 100%, Specificity 83% |  |
| Tooiszadeh et al. 2016 (36) | Validate the accuracy of Upper-Extremity-Frailty (UEF) assessment in distinguishing between F and NF participants | Hospital-based | 101 hospital in-patients aged > 65 years old | Speed of elbow flexion, Number of flexions, Power, and Moment | TSFI | BioSensics LLC: near biceps and wrist | LGR | Sensitivity 78%<br>Specificity 82% | This study shows that a single sensor worn at wrist could be a viable tool for frailty assessment tool however a higher sensitivity would be better on frailty screening use-case. |
| Millor et al. 2017 (37) | Participants performed as many CST reps as possible within 30s at self-selected speed starting from seated position with arms folded | Not specified | A total of 718 subjects from an elderly population aged over 70 years | Temporal-spatial gait parameters: Gait Velocity, Step Regularity, Stride Regularity, Symmetry, Step Time variability | FFP | MTx XSENS worn on L3 spine | Decision Tree: GV | AUC 0.82 | The results showed that the sensors are useful for frailty classification using gait parameters |
| Lee et al. 2018 (38) | Participants wore sensors while performing elbow flexion and extension in 20s timeframe to provide physical frailty assessment features | Hospital-based | N = 100 in-patients (old adults) aged over 70 years old | Mean of angle range, PD of power range, CV of elbow extension time, mean of elbow flexion time, CV of elbow flexion time | Rockwood's criteria (TSFI) | LEGSys worn at wrist and upper arm | LR | AUC 0.87 | This study shows that a single sensor worn at wrist could be a viable tool for frailty assessment tool |

**Abbreviations:** AUC, Area under the Receiver Operating Characteristic curve; BMI, Body Mass Index; CFS, Clinical Frailty Scale; CST, Chair-sit-test; CV, coefficient of variation; EC, eyes closed; EO, eyes open; F, Frailty; FFP, Fried Frailty Phenotype; FTSS, Five Time Sit to Stand; GV, Gait Velocity; LDA, Linear Discriminant Analysis; LGR, Logistic Regression; LR, Linear Regression; NF, Non-frailty; OLCL, open-loop close-loop; PD, percentage of decline; PF, Pre-frail; RF, Random Forest; SVM, Support Vector Machine; TSFI, Rockwood's criteria; TUG, Time-up-and-go

## Clinical Data

Aznar-Tortonda et. al. 2020 collected data from 621 community-based participants in a cross-sectional observational study utilizing an Android mobile device application. Sex, age, polypharmacy, hospitalization, and diabetes history were chosen characteristics and employed in a logistic regression model (32). This model obtained an AUC of 0.78, suggesting that a brief clinical history might be utilized to classify frailty in older persons. Sajeev et. al. 2022 used 20 anthropometric, environmental, social, lifestyle, and physiologic variables from 656 community-dwelling adults aged 40–65 years old to develop and internally validate four machine learning models, including logistic regression, linear discriminant analysis, support vector machine, and random forest (33). With an AUC of 70.8, the random forest model achieved the highest discrimination performance. This study found that machine learning models could be used to diagnose frailty. However, the large number of variables in the purposed models could make it difficult to implement them in clinical practices and community settings, and the selected features appeared to be more difficult to measure and more complicated than the standard diagnostic criteria for frailty. A future study is required to demonstrate the real-world application of a frailty diagnostic machine learning model based on clinical characteristic data.

## Data derived from wearable devices and biological sensors

Most studies for biological sensors and wearable devices employ criteria similar to characteristics research. We divided the parameters into three major categories: 1.) Physical Function test; 2.) Gait and balance test; and 3.) Non-gait-related test.

### Physical Function Test

Greene et. al. (2014) created a support vector machine classifier model based on characteristics gathered from 124 community-dwelling people who wore inertial and pressure sensors on each shin, right thigh, L5 spine, and sternum while undertaking Time-up-and-go, Five Time Sit to Stand, and Balance tests (34). Their model had 88.63% sensitivity and 85.06% specificity, indicating that the demonstrated tests had good frailty classifying characteristics. Schwenk et. al. (2015) had 125 community-dwelling older adults walk 4.57 meters in their home at their own pace, followed by a balance assessment while wearing multiple sensors on their shanks, thighs, and lumbar spine to collect gait and balance parameters for logistic regression model development (39). The results revealed an AUC of 0.857 for non-frail and pre-frail classification and an AUC of 0.841 for pre-frail and frail classification. The mentioned models have shown good and applicable discrimination performance.

### Gait and Balance Test

Tooiszadeh et. al. (2015) demonstrated that postural sway, age, and BMI parameters derived from sensors located at the lumbar spine and shin could predict frailty with 97% sensitivity and 88% specificity (35). Millor et. al. (2017) developed decision tree models using gait characteristics acquired from an inertia sensor worn on the L3 spine of 718 senior volunteers aged over 70 years (37). With an AUC of 0.823-0.896, the results also demonstrated that gait characteristics and decision tree models were beneficial for frailty classification.

### Upper Extremity

According to Lee et. al. 2018, participants wore accelerometers and gyroscope sensors at their wrist and upper arm while performing elbow flexion and extension in a 20-second timeframe to provide physical features such as the mean of the angle range coefficient of variation of elbow flexion and extension time and the mean of elbow movement time (38). These characteristics were used to develop a linear regression model with an AUC of 0.87.

Tooiszadeh et. al. (2016) created a logistic regression model utilizing upper-extremity frailty assessment data from a wearable gyroscope sensor, which was collected from the upper extremities of 101 hospital in-patients over the age of 65 (36). The study’s performance was 78% sensitivity and 82% specificity. These studies demonstrated that a single non-gait-related sensor could be used to distinguish frailty and robustness in the elderly population.

In conclusion, research revealed that physical function tests, gait-related, and non-gait-related measures were useful in developing prediction models to diagnose frailty state in the aged population. However, the fitness test approach may be unsuitable for prospective frailty data collection because performing all the aforementioned fitness tests would take a significant amount of time to obtain the required feature in order to diagnose frailty in an individual, which may be comparable to simply performing tests according to Fried’s criteria. We propose that future research should focus on upper extremity features because we believe that integrating a frailty predictive model into a smartwatch and mobile application has clinically significant implications.

## Therapeutic studies

Based on the current evidence summarized in Table 3, pre-frail and frail older adults are recommended for multi-component physical activity program and progressive resistance training program. Multiple studies have shown improved cognitive function, physical function, and frailty status in older adults after physical exercise intervention. Therefore, our review selected frailty therapeutic studies that integrated the use of technology to improve frailty state in the elderly.

**Table 3.**
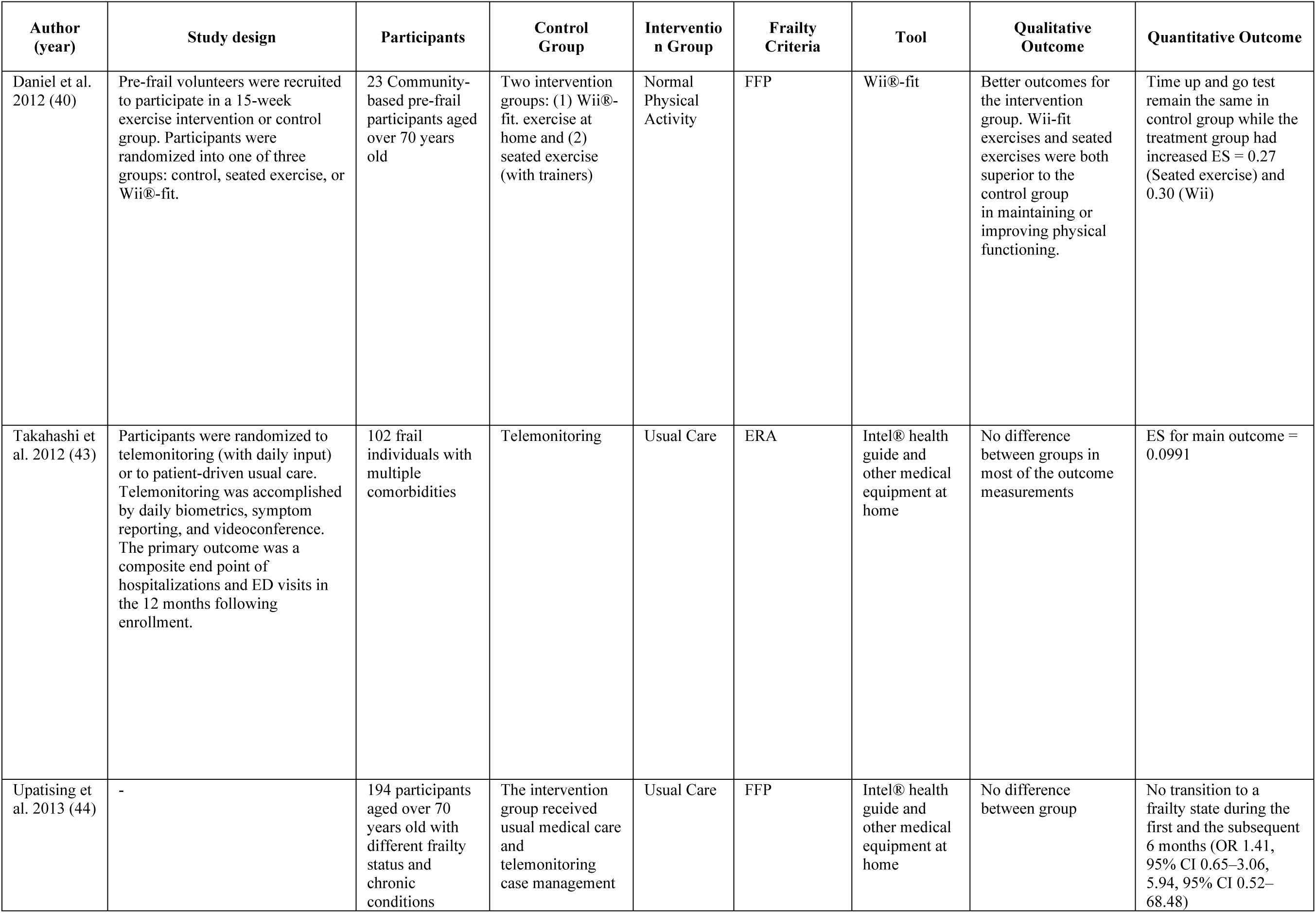
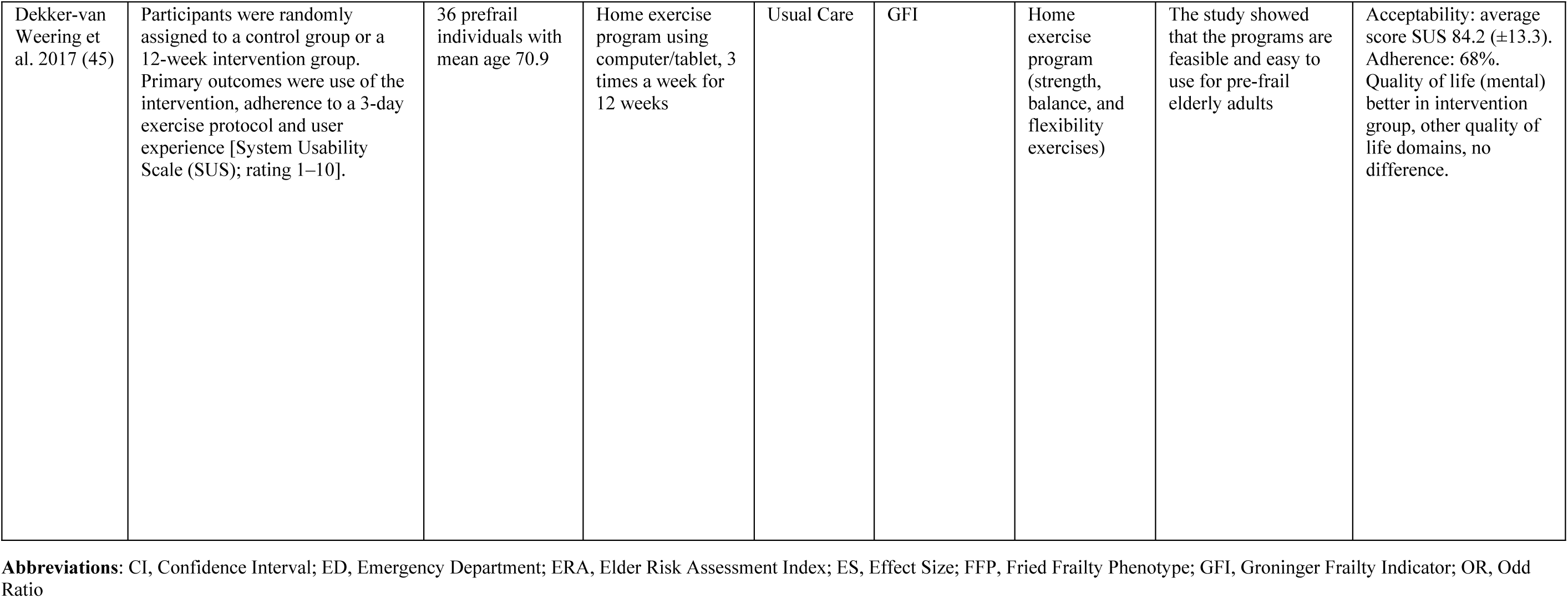
Therapeutic studies.

| Author (year) | Study design | Participants | Control Group | Intervention Group | Frailty Criteria | Tool | Qualitative Outcome | Quantitative Outcome |
| --- | --- | --- | --- | --- | --- | --- | --- | --- |
| Daniel et al. 2012 (40) | Pre-frail volunteers were recruited to participate in a 15-week exercise intervention or control group. Participants were randomized into one of three groups: control, seated exercise, or Wii®-fit. | 23 Community-based pre-frail participants aged over 70 years old | Two intervention groups: (1) Wii®-fit. exercise at home and (2) seated exercise (with trainers) | Normal Physical Activity | FFP | Wii®-fit | Better outcomes for the intervention group. Wii-fit exercises and seated exercises were both superior to the control group in maintaining or improving physical functioning. | Time up and go test remain the same in control group while the treatment group had increased ES = 0.27 (Seated exercise) and 0.30 (Wii) |
| Takahashi et al. 2012 (43) | Participants were randomized to telemonitoring (with daily input) or to patient-driven usual care. Telemonitoring was accomplished by daily biometrics, symptom reporting, and videoconference. The primary outcome was a composite end point of hospitalizations and ED visits in the 12 months following enrollment. | 102 frail individuals with multiple comorbidities | Telemonitoring | Usual Care | ERA | Intel® health guide and other medical equipment at home | No difference between groups in most of the outcome measurements | ES for main outcome = 0.0991 |
| Upatising et al. 2013 (44) | - | 194 participants aged over 70 years old with different frailty status and chronic conditions | The intervention group received usual medical care and telemonitoring case management | Usual Care | FFP | Intel® health guide and other medical equipment at home | No difference between group | No transition to a frailty state during the first and the subsequent 6 months (OR 1.41, 95% CI 0.65–3.06, 5.94, 95% CI 0.52–68.48) |
| Dekker-van Weering et al. 2017 (45) | Participants were randomly assigned to a control group or a 12-week intervention group. Primary outcomes were use of the intervention, adherence to a 3-day exercise protocol and user experience [System Usability Scale (SUS); rating 1–10]. | 36 prefrail individuals with mean age 70.9 | Home exercise program using computer/tablet, 3 times a week for 12 weeks | Usual Care | GFI | Home exercise program (strength, balance, and flexibility exercises) | The study showed that the programs are feasible and easy to use for pre-frail elderly adults | Acceptability: average score SUS 84.2 ($\pm 13.3$). Adherence: 68%. Quality of life (mental) better in intervention group, other quality of life domains, no difference. |

Daniel et. al. 2012 conducted a study where 23 community-dwelling pre-frail volunteers aged over 70 years old were randomized into one of three groups: control, seated exercise, or Wii®-fit. The findings showed better outcomes for all intervention groups (40). Wii-fit exercises and seated exercises were both superior to the control group in maintaining or improving physical functions. Liao et. al. 2019 recruited a randomized controlled trial of 52 prefrail and frail elderly where the participants were divided into two exercise intervention 1.) Exergaming group and 2.) Combined resistance, aerobic and balance exercise group for 36 sessions over 12 weeks (41). The results revealed both gaming exercise and combined exercise groups improved frailty status among the elderly. The study correlated with Moreira et. al. 2021 where an RCT of 66 pre-frail older adults were assigned to either exergaming intervention and traditional multicomponent exercise (42). The findings showed that both programs were clinically effective for delaying frailty status and improving physical and cognitive function.

Exergaming have shown positive health outcomes in terms of enhancing physical function, cognitive function, and frailty status. The programs could be done in a home setting, making exercise intervention easily accessible. However, the majority of frail people are older adults, who may face challenges using technologies because of their lack of digital literacy and technology acceptance. One of the studies cited above had a dropout rate of over 30%, which suggests that a portion of older persons might not find the use of a digital intervention tool appropriate.

## Conclusion

From our review, we found that there are many potential etiognostic factors that could help diagnose frailty status using digital tools from trunk, gait, upper-extremity, and physical activity parameters. Researchers had used these parameters to create multiple well-performing models to classify frailty status in the older adults. We found non-gait parameters the most appealing variables for future research as a frailty diagnostic model integration into a wearable device. However, the model classification results should be interpreted with caution because these models may be overfitting due to a lack of external validation studies.

Regardless of the tools used, studies have shown that exercise can improve frailty status. Rather than developing a single standalone exercise platform, digital health technology developers should focus on how to implement these therapeutic platforms with health care providers or coaching platforms that could encourage and motivate prefrail and frail old adults to engage in more physical activity.

However, there are gaps between frailty screening and care delivery for patient at risk. We did not find a study that combines screening and therapeutic digital health tools into one. To prevent frailty early on, we propose that future studies should focus on how to integrate a screening/diagnostic tool with personalized therapeutic/prevention recommendations to users.

## Data Availability

All relevant data are within the manuscript and its supporting information files.

## Supplementary Materials

None

## Author Contributions

The review’s first draft was written by N.I. and W.S. N.I. and W.S. reviewed and edited the manuscript, assisted in the formulation of the research question, and advised on study objectives and methodologies. N.I. and W.S. contributed to the review’s conceptualization, design, and development. The final version of the manuscript was reviewed and approved by all authors.

## Informed Consent Statement

Not applicable.

## Data Availability Statement

The data presented in this study are available on request from the corresponding author.

## Acknowledgments

This study was partially supported by Chiang Mai University.

## References

1. World population ageing, 2019 : highlights. 2019; Available from: http://digitallibrary.un.org/record/3846855

2. Ageing and health [Internet]. [cited 2022 Aug 2]. Available from: https://www.who.int/news-room/fact-sheets/detail/ageing-and-health

3. Sabri SM, Annuar N, Rahman NLA, Musairah SK, Mutalib HA, Subagja IK. Major Trends in Ageing Population Research: A Bibliometric Analysis from 2001 to 2021. Proceedings. 2022;82(1):19.

4. MacNee W, Rabinovich RA, Choudhury G. Ageing and the border between health and disease. Eur Respir J. 2014 Nov 1;44(5):1332–52.

5. Costa GM da, Sanchez MN, Shimizu HE. Factors associated with mortality of the elderly due to ambulatory care sensitive conditions, between 2008 and 2018, in the Federal District, Brazil. PLOS ONE. 2022 Aug 5;17(8):e0272650.

6. Ayaz T, Sahin SB, Sahin OZ, Bilir O, Rakıcı H. Factors Affecting Mortality in Elderly Patients Hospitalized for Nonmalignant Reasons. J Aging Res. 2014 Aug 3;2014:e584315.

7. Lekan DA, Collins SK, Hayajneh AA. Definitions of Frailty in Qualitative Research: A Qualitative Systematic Review. J Aging Res. 2021 Jun 2;2021:6285058.

8. Sternberg SA, Wershof Schwartz A, Karunananthan S, Bergman H, Mark Clarfield A. The identification of frailty: a systematic literature review. J Am Geriatr Soc. 2011 Nov;59(11):2129–38.

9. Srinonprasert V, Chalermsri C, Aekplakorn W. Frailty index to predict all-cause mortality in Thai community-dwelling older population: A result from a National Health Examination Survey cohort. Arch Gerontol Geriatr. 2018 Jul;77:124–8.

10. End of Life Care in Frailty: Falls [Internet]. British Geriatrics Society. [cited 2022 Aug 2]. Available from: https://www.bgs.org.uk/resources/end-of-life-care-in-frailty-falls

11. Cheng MH, Chang SF. Frailty as a Risk Factor for Falls Among Community Dwelling People: Evidence From a Meta-Analysis: Falls With Frailty. J Nurs Scholarsh. 2017 Sep;49(5):529–36.

12. Davis-Ajami ML, Chang PS, Wu J. Hospital readmission and mortality associations to frailty in hospitalized patients with coronary heart disease. Aging Health Res. 2021 Dec;1(4):100042.

13. Nguyen, Nguyen, Nguyen, Nguyen, Nguyen, Pham, et al. Frailty Prevalence and Association with Health-Related Quality of Life Impairment among Rural Community-Dwelling Older Adults in Vietnam. Int J Environ Res Public Health. 2019 Oct 12;16(20):3869.

14. Robertson DA, Savva GM, Kenny RA. Frailty and cognitive impairment—A review of the evidence and causal mechanisms. Ageing Res Rev. 2013 Sep;12(4):840–51.

15. Castillo-Angeles M, Cooper Z, Jarman MP, Sturgeon D, Salim A, Havens JM. Association of Frailty With Morbidity and Mortality in Emergency General Surgery by Procedural Risk Level. JAMA Surg. 2021 Jan 1;156(1):68–74.

16. Fried LP, Tangen CM, Walston J, Newman AB, Hirsch C, Gottdiener J, et al. Frailty in Older Adults: Evidence for a Phenotype. J Gerontol A Biol Sci Med Sci. 2001 Mar 1;56(3):M146–57.

17. Liu CK, Fielding RA. Exercise as an Intervention for Frailty. Clin Geriatr Med. 2011 Feb;27(1):101–10.

18. Furtado G, Caldo A, Rodrigues R, Pedrosa A, Neves R, Letieri R, et al. Exercise-Based Interventions as a Management of Frailty Syndrome in Older Populations: Design, Strategy, and Planning [Internet]. Frailty in the Elderly - Understanding and Managing Complexity. IntechOpen; 2020 [cited 2022 Dec 20]. Available from: https://www.intechopen.com/state.item.id

19. Fairhall N, Kurrle SE, Sherrington C, Lord SR, Lockwood K, John B, et al. Effectiveness of a multifactorial intervention on preventing development of frailty in pre-frail older people: study protocol for a randomised controlled trial. BMJ Open. 2015 Feb 9;5(2):e007091–e007091.

20. Nakamura M, Ohki M, Mizukoshi R, Takeno I, Tsujita T, Imai R, et al. Effect of Home-Based Training with a Daily Calendar on Preventing Frailty in Community-Dwelling Older People during the COVID-19 Pandemic. Int J Environ Res Public Health. 2022 Oct 30;19(21):14205.

21. Linn N, Goetzinger C, Regnaux JP, Schmitz S, Dessenne C, Fagherazzi G, et al. Digital Health Interventions among People Living with Frailty: A Scoping Review. J Am Med Dir Assoc. 2021 Sep;22(9):1802–1812.e21.

22. Guasti L, Dilaveris P, Mamas MA, Richter D, Christodorescu R, Lumens J, et al. Digital health in older adults for the prevention and management of cardiovascular diseases and frailty. A clinical consensus statement from the ESC Council for Cardiology Practice/Taskforce on Geriatric Cardiology, the ESC Digital Health Committee and the ESC Working Group on e-Cardiology. ESC Heart Fail. 2022;9(5):2808–22.

23. Galán-Mercant A, Cuesta-Vargas AI. Differences in Trunk Accelerometry Between Frail and Nonfrail Elderly Persons in Sit-to-Stand and Stand-to-Sit Transitions Based on a Mobile Inertial Sensor. JMIR Mhealth Uhealth. 2013 Aug 16;1(2):e21.

24. Millor N, Lecumberri P, Gómez M, Martínez-Ramírez A, Izquierdo M. An evaluation of the 30-s chair stand test in older adults: frailty detection based on kinematic parameters from a single inertial unit. J NeuroEngineering Rehabil. 2013 Dec;10(1):86.

25. Toosizadeh N, Mohler J, Najafi B. Assessing Upper Extremity Motion: An Innovative Method to Identify Frailty. J Am Geriatr Soc. 2015 Jun;63(6):1181–6.

26. Parvaneh S, Mohler J, Toosizadeh N, Grewal GS, Najafi B. Postural Transitions during Activities of Daily Living Could Identify Frailty Status: Application of Wearable Technology to Identify Frailty during Unsupervised Condition. Gerontology. 2017;63(5):479–87.

27. Castaneda-Gameros D, Redwood S, Thompson JL. Physical Activity, Sedentary Time, and Frailty in Older Migrant Women From Ethnically Diverse Backgrounds: A Mixed-Methods Study. J Aging Phys Act. 2018 Apr;26(2):194–203.

28. Zhou H, Razjouyan J, Halder D, Naik AD, Kunik ME, Najafi B. Instrumented Trail-Making Task: Application of Wearable Sensor to Determine Physical Frailty Phenotypes. Gerontology. 2019;65(2):186–97.

29. Jansen CP, Toosizadeh N, Mohler MJ, Najafi B, Wendel C, Schwenk M. The association between motor capacity and mobility performance: frailty as a moderator. Eur Rev Aging Phys Act. 2019 Dec;16(1):16.

30. Apsega A, Petrauskas L, Alekna V, Daunoraviciene K, Sevcenko V, Mastaviciute A, et al. Wearable Sensors Technology as a Tool for Discriminating Frailty Levels During Instrumented Gait Analysis. Appl Sci. 2020 Nov 26;10(23):8451.

31. Kikuchi H, Inoue S, Amagasa S, Fukushima N, Machida M, Murayama H, et al. Associations of older adults’ physical activity and bout-specific sedentary time with frailty status: Compositional analyses from the NEIGE study. Exp Gerontol. 2021 Jan;143:111149.

32. Aznar-Tortonda V, Palazón-Bru A, la Rosa DMF de, Espínola-Morel V, Pérez-Pérez BF, León-Ruiz AB, et al. Detection of frailty in older patients using a mobile app: cross-sectional observational study in primary care. Br J Gen Pract J R Coll Gen Pract. 2020 Jan;70(690):e29–35.

33. Sajeev S, Champion S, Maeder A, Gordon S. Machine learning models for identifying pre-frailty in community dwelling older adults. BMC Geriatr. 2022 Oct 12;22(1):794.

34. Greene BR, Doheny EP, Kenny RA, Caulfield B. Classification of frailty and falls history using a combination of sensor-based mobility assessments. Physiol Meas. 2014 Oct 1;35(10):2053–66.

35. Toosizadeh N, Mohler J, Wendel C, Najafi B. Influences of Frailty Syndrome on Open-Loop and Closed-Loop Postural Control Strategy. Gerontology. 2015;61(1):51–60.

36. Toosizadeh N, Joseph B, Heusser MR, Orouji Jokar T, Mohler J, Phelan HA, et al. Assessing Upper-Extremity Motion: An Innovative, Objective Method to Identify Frailty in Older Bed-Bound Trauma Patients. J Am Coll Surg. 2016 Aug;223(2):240–8.

37. Millor N, Lecumberri P, Gomez M, Martinez A, Martinikorena J, Rodriguez-Manas L, et al. Gait Velocity and Chair Sit-Stand-Sit Performance Improves Current Frailty-Status Identification. IEEE Trans Neural Syst Rehabil Eng. 2017 Nov;25(11):2018–25.

38. Lee H, Joseph B, Enriquez A, Najafi B. Toward Using a Smartwatch to Monitor Frailty in a Hospital Setting: Using a Single Wrist-Wearable Sensor to Assess Frailty in Bedbound Inpatients. Gerontology. 2018;64(4):389– 400.

39. Schwenk M, Mohler J, Wendel C, D’’Huyvetter K, Fain M, Taylor-Piliae R, et al. Wearable Sensor-Based In-Home Assessment of Gait, Balance, and Physical Activity for Discrimination of Frailty Status: Baseline Results of the Arizona Frailty Cohort Study. Gerontology. 2015;61(3):258–67.

40. Daniel K. Wii-Hab for Pre-Frail Older Adults. Rehabil Nurs. 2012 Jul;37(4):195–201.

41. Liao YY, Chen IH, Wang RY. Effects of Kinect-based exergaming on frailty status and physical performance in prefrail and frail elderly: A randomized controlled trial. Sci Rep. 2019 Jun 27;9(1):9353.

42. Moreira NB, Rodacki ALF, Costa SN, Pitta A, Bento PCB. Perceptive-Cognitive and Physical Function in Prefrail Older Adults: Exergaming Versus Traditional Multicomponent Training. Rejuvenation Res. 2021 Feb;24(1):28–36.

43. Takahashi PY, Pecina JL, Upatising B, Chaudhry R, Shah ND, Van Houten H, et al. A Randomized Controlled Trial of Telemonitoring in Older Adults With Multiple Health Issues to Prevent Hospitalizations and Emergency Department Visits. Arch Intern Med [Internet]. 2012 May 28 [cited 2022 Dec 20];172(10). Available from: http://archinte.jamanetwork.com/article.aspx?doi=10.1001/archinternmed.2012.256

44. Upatising B, Hanson, Kim, Cha, Yih Y, Takahashi P. Effects of home telemonitoring on transitions between frailty states and death for older adults: a randomized controlled trial. Int J Gen Med. 2013 Mar;145.

45. Dekker-van Weering M, Jansen-Kosterink S, Frazer S, Vollenbroek-Hutten M. User Experience, Actual Use, and Effectiveness of an Information Communication Technology-Supported Home Exercise Program for Pre-Frail Older Adults. Front Med. 2017 Nov 27;4:208.

